# Thrombin generation is associated with extracellular vesicle and leukocyte lipid membranes in acute coronary syndrome

**DOI:** 10.1101/2023.05.03.23289247

**Authors:** Majd B Protty, Victoria J Tyrrell, Keith Allen-Redpath, Shin Soyama, Ali A Hajeyah, Daniela Costa, Anirban Choudhury, Rito Mitra, Parveen Yaqoob, P Vince Jenkins, Zaheer Yousef, Peter W Collins, Valerie B O’Donnell

## Abstract

**Background:** Acute coronary syndrome (ACS) is caused by arterial thrombosis and is associated with sustained activation of coagulation. Clotting requires interactions of coagulation factors with aminophospholipids (aPL): phosphatidylserine (PS) and phosphatidylethanolamine (PE) on membrane surfaces. The aPL composition of circulating membranes in coronary disease has not been characterized. Furthermore, the contribution of external-facing aPL to elevated thrombotic risk in ACS is unknown.

**Methods and results:** Thrombin generation was measured on platelet, leukocyte and extracellular vesicles (EV) from patients with ACS (n = 24), stable coronary artery disease (CAD, n = 18), risk factor positive (RF, n = 23) and healthy controls (HC, n = 24). The aPL composition on the surface of EV, platelets and leukocytes was determined using lipidomics. Leukocytes, platelets and EV externalized PE- and PS-containing fatty acids ranging from C16:0-20:4. These included both diacyl and plasmalogen forms, with significant increases stimulated by agonist activation. Thrombin generation on the surface of EV and leukocytes was higher in ACS than HC. Also, thrombin generation was higher for EV from CAD and RF, than HC. EV counts were higher in CAD and ACS compared with HC. Thrombin generation correlated positively with plasma EV counts and membrane surface area.

**Conclusion:** The aPL membrane of EV and leukocytes may contribute to the activation of coagulation in CAD and ACS. Targeting EV formation/clearance and the aPL surface of EV and leukocyte membranes represents a novel anti-thrombotic target in CAD and ACS.

**Condensed abstract:** Acute coronary syndrome (ACS) is associated with sustained activation of coagulation, requiring procoagulant aminophospholipids (aPL). However, the aPL composition of circulating membranes and their contribution to thrombotic risk in ACS is undetermined. Lipidomics demonstrated that leukocytes, platelets and extracellular vesicles (EV) externalized aPL-containing fatty acids ranging from C16:0-20:4. Thrombin generation on the surface of EV and leukocytes was higher in ACS patients than healthy controls (HC). EV counts were higher in ACS compared with HC and correlated positively with thrombin generation. In summary, aPL in the outer membranes of EV and leukocytes may contribute to elevated thrombotic risk in ACS.

**Highlights:** *What is new?:*

- The aPL profile of platelets, leukocytes and EV in patients with ACS, CAD, RF and HC is defined for the first-time using lipidomics.
- Thrombin generation on the surface of unstimulated leukocytes, is elevated in patients with ACS compared with HC.
- Thrombin generation on the surface of EV is elevated in patients with ACS, CAD and RF compared with HC.
- EV counts in patients with ACS/CAD/RF were elevated compared with HC and correlate positively with thrombin generation.

**Clinical Perspective:** *What are the clinical implications?:*

- The membranes of EV and leukocytes may contribute to the activation of coagulation in ACS.
- The aPL in EV and leukocyte membranes represent a novel target for reducing thrombotic risk in ACS.
- Targeting EV formation/clearance could reduce thrombotic risk in CAD and ACS.

## Introduction

Acute coronary syndrome (ACS) is the commonest cause of ischemic heart disease and is associated with high rates of mortality, recurrent infarction and other complications^1–5^. It affects 150,000 people per year (incidence) in the United Kingdom, incurring £3.6 billion of annual direct healthcare costs^6, 7^. The underlying pathophysiology is an inflammatory process sparked by atherosclerotic plaque rupture leading to activation and recruitment of platelets and leukocytes along with upregulation of tissue factor (TF) expression^8–15^. This activates coagulation, forming an occlusive arterial thrombus driving ischemia and infarction^16^. Despite standard treatment with anti-platelet agents, the rates of subsequent strokes, myocardial infarction and cardiovascular death exceed 10% in the first year post ACS diagnosis^17, 18^. In line with this, patients with ACS have elevated and sustained plasma thrombin generation for months beyond the acute insult as evidenced by higher levels of plasma thrombin:anti-thrombin complexes (TAT)^19, 20^. This indicates that there are likely to be other modifiable factors involved beyond platelet activity^17, 18, 21^.

Thrombin generation requires a procoagulant phospholipid (PL) membrane to allow the assembly of the “prothrombinase” (Factor Xa/Va) complex and other Gla-domain containing coagulation factors^22^. This can be provided by the external surface of activated platelets, leukocytes and extracellular vesicles (EV)^23, 24^. Resting platelet and leukocyte membranes are comprised primarily of phosphatidylcholine (PC) on the external side, with the aminophospholipids (aPL) phosphatidylethanolamine (PE) and phosphatidylserine (PS) being internally facing^25^. The binding of coagulation factors requires a specific phospholipid (PL) composition which includes the electronegative headgroup of PS, supported by PE, and is dependent on the presence of calcium ions^26, 27^. Native PC does not facilitate the binding of coagulation factors and so resting platelets or leukocytes provide minimal support for coagulation reactions. During inflammation or acute trauma/bleeding challenge, platelets and leukocytes become activated, leading to a calcium-dependent translocation of PS/PE to the outside of the cell, mediated by scramblase^28^. Inflammation also leads to the release of aPL-rich EV. Together, this exposure of aPL to the circulation leads to interactions with coagulation factors via calcium which are essential for effective coagulation in vivo^29–31^.

Whether the enhanced thrombotic risk that persists following ACS is related to altered levels of aPL-mediated thrombin generation on the surface of blood cells is currently unknown. Previous studies have focused on differences in coagulation factor amounts and activity, but there have been no studies focusing on the contribution of the PL membrane to thrombin generation independent of tissue factor or plasma content^32^. To determine this, we characterized the aPL membrane composition in platelets, leukocytes and EV in patients with ACS, stable coronary artery disease (CAD) or risk factors thereof (RF). We assessed the ability of these membranes to support coagulation in vitro compared with healthy controls (HC) using a thrombin generation assay that employs purified coagulation factors independent of tissue factor^33–36^. This approach allowed the contribution of the pro-coagulant membrane to coagulation to be determined specifically.

## Methods

### Study Participants for clinical cohort (blood samples)

#### Clinical cohort

Participants were recruited from Cardiff University and Cardiff and Vale University Health Boards. Ethical approval was from Health and Care Research Wales (HCRW, IRAS 243701; REC reference 18/YH/0502). Age and sex-matched individuals were recruited into one of the following four groups: *(i) Acute Coronary Syndrome (ACS)*: Participants were identified on in-patient cardiology wards using diagnostic tests (ischemic ECG changes, raised troponin level above normal laboratory defined range) and clinical assessment by the cardiology team. All were recruited within 48 hours of the index event prior to any revascularisation/angioplasty. *(ii) Significant coronary artery disease (CAD):* Patients attending for an elective coronary angiogram to assess for symptoms of stable angina in the absence of a history of ACS. Coronary angiography demonstrated lesions requiring revascularization on anatomical/physiological criteria as defined by guidelines from the European society for cardiology (ESC, 2018)^37^. *(iii) Risk-factor controls with no significant CAD (RF):* This group includes patients attending for a diagnostic coronary angiogram with risk factors for ischemic heart disease (a clinical diagnosis of hypertension requiring therapy, diabetes types 1 or 2, hypercholesterolemia [total cholesterol > 6 mmol/L], smoking, chronic kidney disease stage 3 or more, or combination thereof) but whose coronary angiogram demonstrates no significant coronary artery disease, defined as not requiring revascularization on anatomical/physiological criteria as per the ESC 2018 guidelines^37^. *(iv) Healthy controls (HC):* Participants had no significant history for ischemic heart disease or its risk factors, were never-smokers, and were not on anti-platelet agents, anti-coagulants or statins. They were identified from the workplace or were volunteers from partner studies such as ‘HealthWise Wales’^38^. Clinical characteristics are in Table 1. Inclusion criteria were aged 18 years or over, acute coronary syndrome in ACS group, and no history of ACS in the others. Exclusion criteria were: diagnosis of infective endocarditis or atrial fibrillation, or inability to consent to study. Overall, 90 participants were recruited: HC, n = 24, RF, n = 23, CAD, n = 19, ACS, n = 24. Blood samples were collected by peripheral venepuncture as outlined below, by one individual and all samples were transferred to the laboratory within 10 min. The study design is summarized in **Supplementary Figure 1**.

### Platelet isolation

Whole blood was taken from using a 21G butterfly needle into a 50 ml syringe containing acidified citrate dextrose (ACD; 85 mM trisodium citrate, 65 mM citric acid, 100 mM glucose) at a ratio of 8.1 parts whole blood to 1.9 parts ACD, as described previously^33^, and centrifuged at 250 *g* for 10 min at 20 °C. The platelet-rich plasma was collected and centrifuged at 1000 *g* for 8 min at 20 °C. Platelet poor plasma was removed and retained for extracellular vesicle isolation. The platelet pellet was resuspended in Tyrode’s buffer (134 mm NaCl, 12 mm NaHCO_3_, 2.9 mm KCl, 0.34 mm Na_2_HPO_4_, 1.0 mm MgCl_2_, 10 mm Hepes, 5 mm glucose, pH 7.4) containing ACD (9:1, v/v). The platelets were washed by centrifuging at 1000 *g* for 8 min at 20 °C then resuspended in Tyrode’s buffer at 2 × 10^8^·ml^−1^. Platelets were activated at 37 °C in the presence of 1 mM CaCl_2_, 0.2 unit·ml^−1^ thrombin (Sigma Aldrich).

### Leukocyte isolation

Leukocytes were isolated from 20 ml citrate-anticoagulated whole blood as described previously^33^. Briefly, 20 ml of blood was drawn using a 21G butterfly needle into a 50 ml syringe containing 4 ml of 2 % citrate and 4 ml of Hetasep (Stem Cell Technologies) and allowed to sediment for 45 minutes. The upper plasma layer was recovered and centrifuged at 250 g for 10 min at 4 °C. The pellet was resuspended in ice-cold 0.4 % trisodium citrate/PBS and centrifuged at 250 g for 5 min at 4 °C. Erythrocytes were removed by hypotonic lysis (0.2 % hypotonic saline before being neutralized with a PBS wash. Leukocytes were resuspended in Krebs buffer (100 mM NaCl, 48 mM HEPES, 5 mM KCl, 1 mM sodium dihydrogen orthophosphate dihydrate and 2 mM glucose) at 4 x 10^6^/ml. For activation, 4 x 10^6^ leukocytes were incubated at 37 °C with 10 μM A23187 and 1 mM CaCl_2_, for 30 min, prior to lipid extraction.

### Extracellular vesicle (EV) isolation

Methods were adapted from recent literature and guidelines^39, 40^. Platelet poor plasma generated as above was centrifuged at 1000 g for 10 min at 20 °C to generate platelet-free plasma (PFP). 1 ml PFP was snap frozen on dry ice and stored at −80 °C for quantification at a later date as below. For each donor’s plasma, 6 x 1 ml PFP aliquots were centrifuged at 16,000 g for 30 min at 20 °C. 750 μl was removed from each aliquot, and 750 μl of modified Tyrode’s buffer was added to the pellet, which was gently resuspended using a pipette. Following a second centrifugation as above, 950 μl was removed. 50 μl modified Tyrode’s buffer was added to the pellet to gently resuspend and recover the EV-rich fraction. EV fractions were pooled to generate one isolate per donor. Of this, 250 μl was used for lipidomics, and 3 x 20 μl for prothrombinase assays.

### Extracellular vesicle (EV) quantification

EV quantification was performed by thawing one aliquot of PFP per patient, of which 500 μL was passed through size-exclusion chromatography iZON qEV columns (Izon Science Ltd, UK) to recover particles and vesicles between 70 nm to 1000 nm in diameter. The eluting EV-rich fractions were collected and analyzed using nanoparticle tracking on a NanoSight 300 (Malvern, UK) equipped with a sensitive sCMOS camera and a 488 nm blue laser, to generate an EV count and size distribution for all participants.

### Prothrombinase assay

Resting platelets (4 x 10^6^), resting leukocytes (8 x 10^4^) and plasma EV isolates (20 μL) were added in triplicate to a 96-well half-area flat bottom clear plate (Greiner, Austria). Next, a mix of recombinant factor Xa (FXa) (50 nM, Enzyme Research Laboratories, UK), factor Va (FVa) (15 nM, Haematologic/Cambridge Bioscience, UK), factor II (FII) (1 μM, Enzyme Research Laboratories, UK) and CaCl_2_ (5 mM) in prothrombinase buffer (20 mM Tris, 150 mM NaCl, 0.05% BSA w/v) was added to the wells and the reaction allowed to proceed for 5 min at 21 °C, before being quenched with an excess of EDTA (7 mM final concentration). Thrombin (FIIa) activity was measured on a plate reader using a chromogenic substrate as described in Supplementary Methods. The assay is depicted graphically in **Supplementary Figure 2**.

### Lipid biotinylation, extraction and analysis for aPL

To determine the amounts of PE and PS on the external leaflet of cell membranes, total and external aPL were measured as described previously^41^. Briefly, 0.2 mL of platelets (4 x 10^7^), leukocytes (8 x 10^5^) or EV isolated as described above were incubated with 20 μL of 20mM NHS-biotin for 10 min at 21 °C to label total aPL. In the case of externalized aPL, samples were incubated with 86 μL of 11mM EZ-Link sulfo-NHS-biotin for 10 min at 21 °C followed by 72 μl of 250 mM of L-lysine for 10 min at 21 °C. The final volumes were made up to 0.4 mL with phosphate-buffered saline (PBS). Lipids were extracted by adding samples to 1.5 mL chloroform:methanol (1:2) containing 10 ng internal standards (biotinylated 1,2- dimyristoyl-PE and -PS, generated as in Thomas et al^41^) to give a solvent:sample ratio of 3.75:1, as described previously^41^. Following vortexing and centrifugation (400 g, 5 mins), lipids were recovered in the lower chloroform layer, which was dried under vacuum. Samples were analyzed for aPL using LC-MS/MS. For this, samples were separated on an Ascentis C-18 (5 μm 150 mm × 2.1 mm) column (Sigma Aldrich, USA) with an isocratic gradient (methanol, 0.2 % w/v NH_4_CH_3_CO_2_) at a flow rate of 400 μL/min. Products were analyzed in MRM mode on a Q-Trap 4000 instrument (Applied Biosystems, UK) by monitoring precursor-to-product ion transitions in negative ion mode (**Supplementary Table 2**). The peak area for the analytes was integrated and normalized to the internal standards. Limit of quantitation (LOQ) is defined as signal:noise of 5:1 with at least 6 data points across a peak. For quantification, standard curves were generated using biotinylated PS and PE^41^. Chromatograms of biotinylated PS and PE species as detected in representative participant samples can be seen in **Supplementary Figures 3 and 4**, respectively.

### Statistical analysis

Statistical significance was determined with the Mann-Whitney-Wilcoxon Test or T-test for pairwise comparison. Correlation analysis utilized Pearson’s correlation for linear dependence between variables. The cut-off value chosen for test significance was a p-value <0.05. Box plots were drawn in Excel (Microsoft, USA) with edges indicating the interquartile range (IQR), the line inside the box indicating the median, and whiskers indicating 1.5 times the IQR. For generation of heatmaps, samples were averaged within their groups, and a log10 was applied to lipid amounts (ng) normalized to cell count or tissue weight (mg) for each lipid to allow row-wise and column-wise comparison. Next, lipid measurements were plotted as intensity values using the ‘pheatmap’ package in the R coding environment (v3.6.2, open source) with lipid hierarchical clustering. Intensity levels were represented by a colour gradient ranging from blue (very low levels or absent) to red (high levels) with variations in between. Graphical illustrations of assays, designs and pathways were carried out using the online platforms draw.io and biorender.com (premium subscription).

## Results

### EV-containing plasma from ACS patients supports elevated thrombin generation, driven by higher EV counts in disease

In an in vitro system using purified FXa, FVa and FII, EV-containing plasma from patients with ACS, CAD or RF stimulated significantly higher thrombin generation than those from HC (**Figure 1 A**). EV were isolated from a fixed volume (6 mL) of plasma, thus, particles were quantified using nanoparticle tracking analysis. There was a trend for higher EV counts in plasma from all patient groups compared with HC, which was significantly higher for CAD (**Figure 1 B**). Furthermore, there was a weak but significant positive correlation between thrombin generation and EV counts (**Figure 1 C**). The mean EV vesicle diameter was smaller in disease groups (RF, CAD and ACS) compared with HC (**Supplementary Figure 5**). As coagulation takes place on the surface of vesicles, we next calculated the EV total surface area (vesicle area as a sphere calculated as 4πr^2^ x EV counts) in plasma. A trend was seen for higher EV total surface areas in patient groups, which was significant for CAD vs HC (**Figure 1 D**). Overall, this indicates that EV from patients tend to be smaller, but due to higher counts, the overall EV surface area will be elevated in disease. This may directly contribute to the elevated thrombin generation observed. Confirming this idea, we normalized thrombin generation to either EV counts or surface area and found that this abolished the differences between groups (**Figure 1 E-F**).

**Figure 1.**
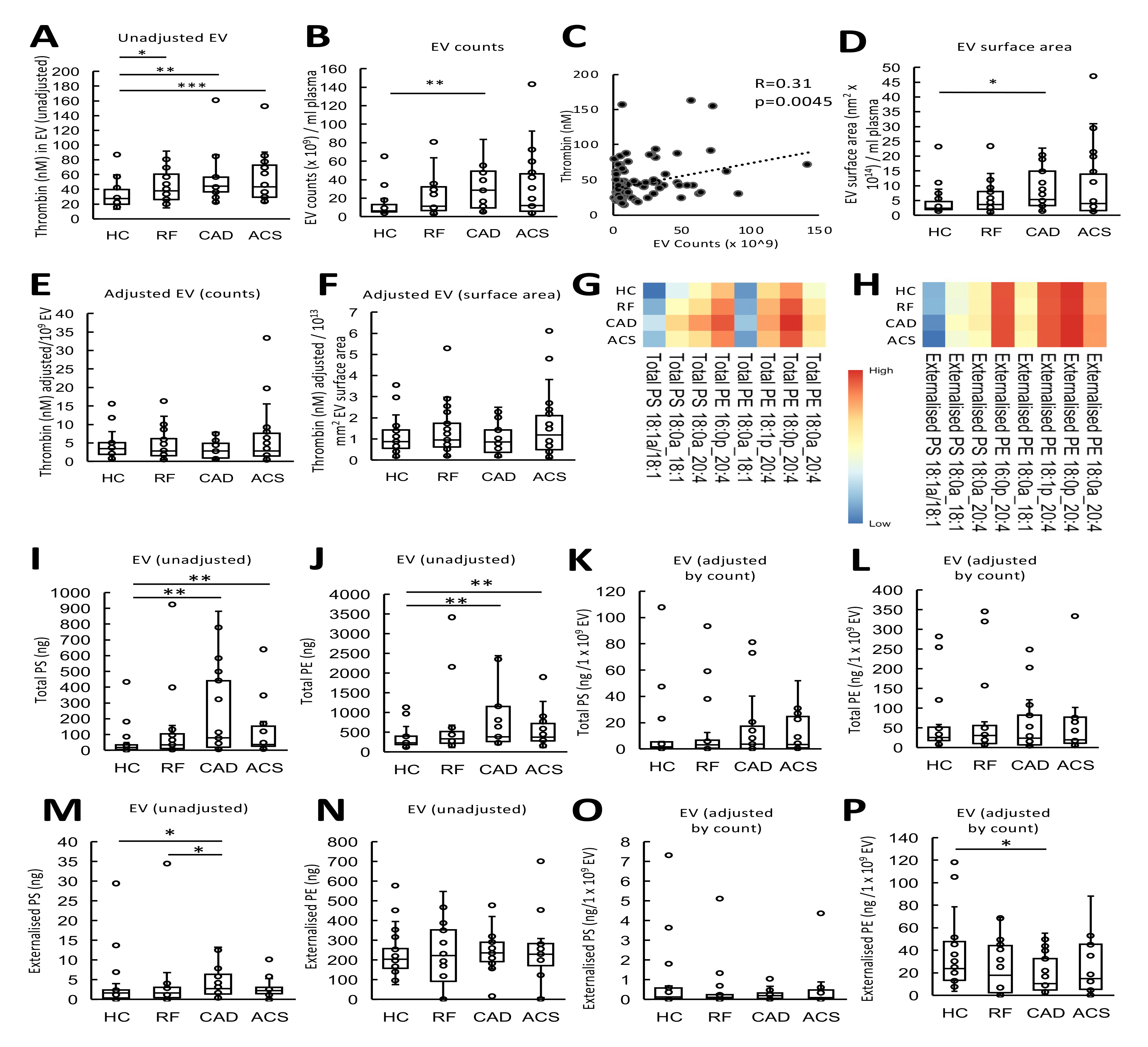
Plasma EV support elevated thrombin generation in coronary artery disease. *Panel A. EV from patients support higher levels of thrombin generation.* The ability of EV membranes to support thrombin generation was assessed using prothrombinase assay as described in Methods. *Panel B. EV counts are significantly higher in patients.* To quantify EV, platelet-free plasma (0.5 mL) from each participant was processed through size-exclusion chromatography (iZON qEV columns) and nanoparticle tracking analysis (Nanosight 300). *Panel C. Thrombin generation positively correlates with EV counts.* Thrombin generation and EV counts were correlated using Pearson’s correlation. *Panel D. EV surface area is increased in vascular disease.* EV surface area was calculated as described in Methods and plotted as box plots. *Panel E,F. Thrombin generation was not changed between groups if EV were normalized by counts or surface area*. Thrombin generation on the surface of EV was adjusted by EV counts (per 1× 10^9^ EV) (Panel E) and surface area (Panel F) and plotted as box plots. *Panels G,H. Heatmaps showing aPL molecular species in EV.* Heatmaps were drawn using the pheatmap R package as described in methods to visualize aPL amounts between groups for all the measured species, analyzed using LC-MS/MS. *Panels I,J. Grouping by headgroup, the amounts of total PS and total PE for each of the clinical groups were plotted to examine for differences between groups. Panel K-L. Total aPL amounts adjusted by EV count. Panel M-N. Grouping by headgroup for externalized PS and PE lipids. Panel O-P. Externalized aPL amounts adjusted by EV count.* Lipids were extracted from EV-rich plasma fractions as in Materials and Methods. Lipids amounts (ng) were calculated by LC-MS/MS. Statistical significance was tested with Mann-Whitney-Wilcoxon test (*: p <0.05, **: p <0.01, ***: p <0.001). ACS: acute coronary syndrome (n=24), CAD: coronary artery disease but no ACS (n=19), RF: Risk factors with no significant coronary artery disease (n=23), HC: Healthy control (n=24).

### Higher amounts of pro-coagulant PE and PS are detected in EV from patient groups

The aPL composition of EV has not been described before, hence total and external PS and PE species were next characterized using LC-MS/MS. In this assay, external facing aPL are derivatized using the cell impermeable biotinylation reagent, sulfo-NHS-biotin. Total aPL is instead derivatized using the cell-permeable form, NHS-biotin^41^. Derivatized aPL are then detected using LC/MS/MS, based on a mass shift of +226 a.m.u. from the native lipid. This method was previously used to determine the molecular species and amounts of PS and PE on the surface of platelets from healthy donors^34^. The most abundant species detected were: PE 16:0p_20:4, PE 18:0a_20:4, PE 18:0p_20:4, PS 18:0a_18:1 and PS 18:0a_20:4 (**Figure 1 G**, Supplementary Figure 6), as previously shown in healthy platelets^34^. Externalized PS and PE comprised the same molecular species, with the most abundant isomers being detected in higher amounts on the outside (**Figure 1 H, Supplementary Figure 6**). This indicates an absence of selectivity for any particular aPL isomers to be present on the outer side of EV.

EV from plasma of ACS and CAD patients contained significantly higher levels of total PS and total PE compared with HC (**Figure 1 I,J**), however, this difference disappeared once EV counts were taken into account (**Figure 1 K,L**). Higher levels of externalized PS were also detected in CAD patients compared with HC/RF, although externalized PE was similar (**Figure 1 M,N**). Again, once adjusted by EV counts, externalized PS/PE levels were similar between groups (**Figure 1 O,P**). As for thrombin generation, this indicates that both total and external facing PE/PS are higher in disease, but this is due to the elevated circulating EV counts in plasma from patient groups.

### Leukocytes from ACS patients support higher levels of thrombin generation than those from HC

Next, we characterized the pro-coagulant membrane on circulating white cells from patient groups, also comparing their ability to support thrombin generation with PE/PS content. Resting leukocytes (8 x 10^4^) from ACS patients supported significantly higher thrombin generation than HC, with a trend for higher thrombin generation in all disease groups (**Figure 2 A**). In the patient groups, there was a trend towards higher circulating leukocyte counts as measured by the hospital clinical laboratory, with worsening disease, although all cell counts were within normal range (**Supplementary Table 1**). To test the potential impact of cell count *in vivo*, we normalized thrombin generation by circulating leukocyte count, since a higher count in disease could further impact thrombin generation. This further demonstrated an upward trend with a significantly higher amount of thrombin generated in ACS compared with RF (**Figure 2 B**). Leukocyte counts had not been obtained for HC samples, as volunteers were outside the hospital system, so direct comparison with this group was not possible.

**Figure 2.**
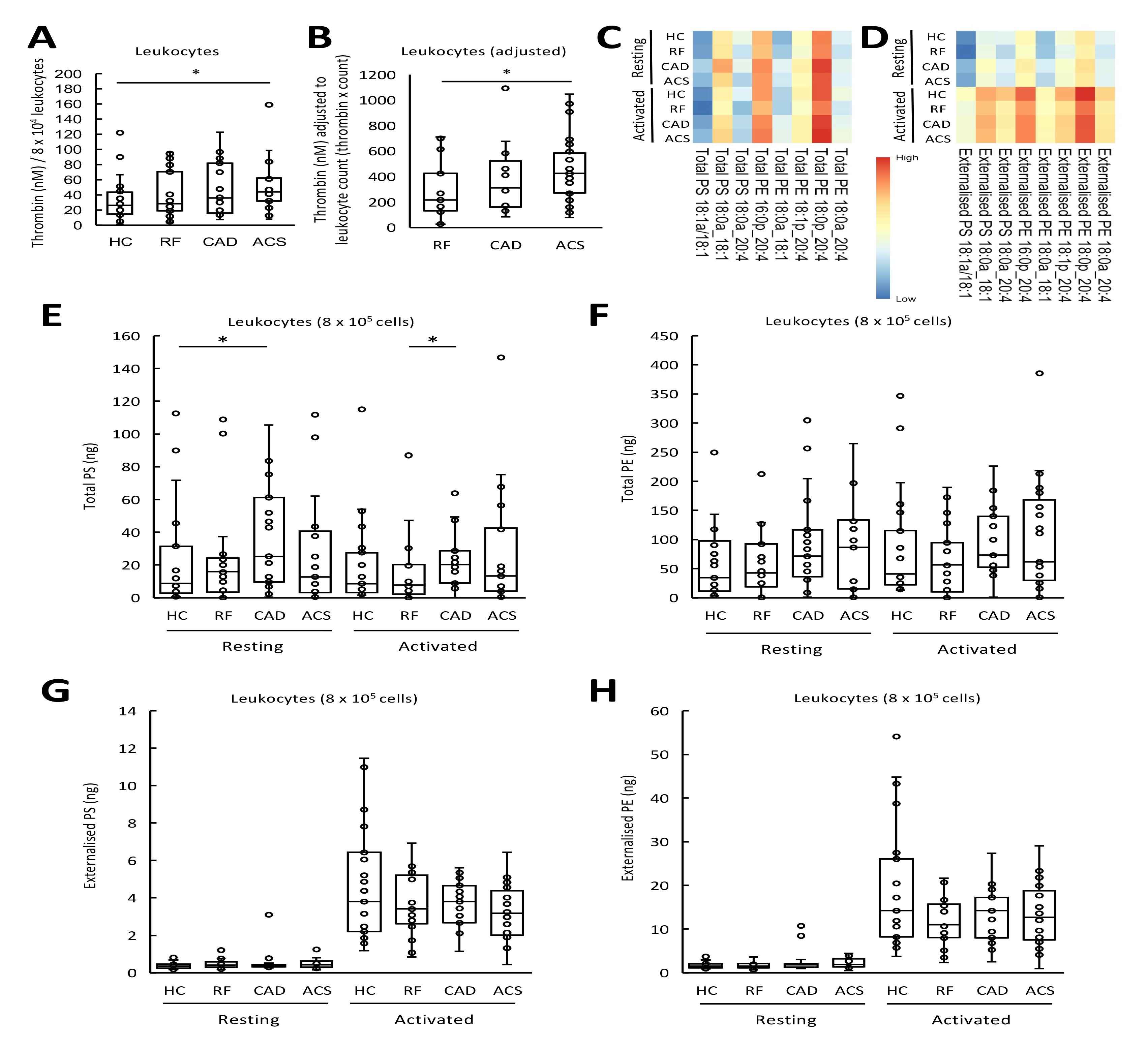
Leukocytes from ACS patients generated more thrombin than HC, and this may be further influenced by higher *in-vivo* leukocyte counts. *Panel A. Leukocytes from ACS patients support higher levels of thrombin generation than HC.* The ability of leukocyte membranes to support thrombin generation was quantified using prothrombinase assay as described in Methods and displayed on a box plot. *Panel B. Adjusting by total leukocyte count demonstrates an upward trend in thrombin generation between RF, CAD and ACS samples.* Thrombin generation was adjusted by total leukocyte count to account for differences between groups. *Panels C,D. Heatmaps show aPL molecular species in leukocytes, with increased externalization upon activation.* Lipids were extracted from resting or ionophore activated leukocytes and quantified using LC-MS/MS as described in Methods. The log10 lipid amounts (ng) were plotted on a heatmap using the pheatmap R package as described in Methods to show total (Panel C) and externalized (Panel D) aPL molecular species. *Panels E-H. Grouping by headgroup, the amounts of total PS, total PE, externalized PS and externalized PE were largely similar between clinical groups.* Lipids were extracted from resting or ionophore-activated leukocytes as described in Methods. Lipids amounts (ng) were determined using LC-MS/MS. Statistical significance was tested with Mann-Whitney-Wilcoxon test (*: p <0.05, **: p <0.01, ***: p <0.001). ACS: acute coronary syndrome (n=24), CAD: coronary artery disease but no ACS (n=19), RF: Risk factors with no significant coronary artery disease (n=23), HC: Healthy control (n=24).

### Leukocytes externalize aPL on activation, while molecular composition is unchanged in coronary artery disease

The aPL composition of leukocyte membranes is currently unknown. To characterize this, and determine the impact of coronary artery disease, total and external molecular species of aPL were quantified in leukocytes from HC and ACS, CAD and RF patients, both before after calcium ionophore activation. Ionophore activation reveals the potential of the cells, when fully activated, to externalize aPL, while resting cells are more comparable with circulating cells. Three species of PS and five of PE were detected, with all PE containing 20:4, showing a similar composition to EV (**Figure 2 C,D, Supplementary Figure 7**). Total PE and PS was similar for all groups, and this was not impacted by activation (**Figure 2 E,F**). In contrast, following ionophore activation, leukocytes from all groups externalized aPL (p<0.001, **Figure 2 G,H**). As for EV, the most abundant isomers were detected in higher amounts on the outside indicating a lack of selectivity for any specific aPL isomer to be externalized (**Figure 2 G,H**). Externalized aPL species or amounts did not differ significantly between patient groups, and there was no correlation between the amount of thrombin generated on the surface of resting leukocytes and externalized PS and PE amounts (**Figure 2, G,H, Supplementary Figure 8**).

These data indicate that the elevated thrombin generation on the surface of leukocytes in ACS compared with HC is unlikely to be related to levels of externalized aPL.

### Thrombin generation on the surface of resting platelets was similar for all groups

Next, the ability of washed platelets from patient groups to support prothrombinase activity was tested. Thrombin generation was strongly stimulated by platelets from all groups tested, but there were no significant differences seen (**Figure 3 A**). Furthermore, platelet counts were similar between disease groups with all being within the normal range. This suggests that platelet membrane support for thrombin generation is not responsible for increased thrombotic risk in ACS. (**Supplementary Table 1**).

**Figure 3.**
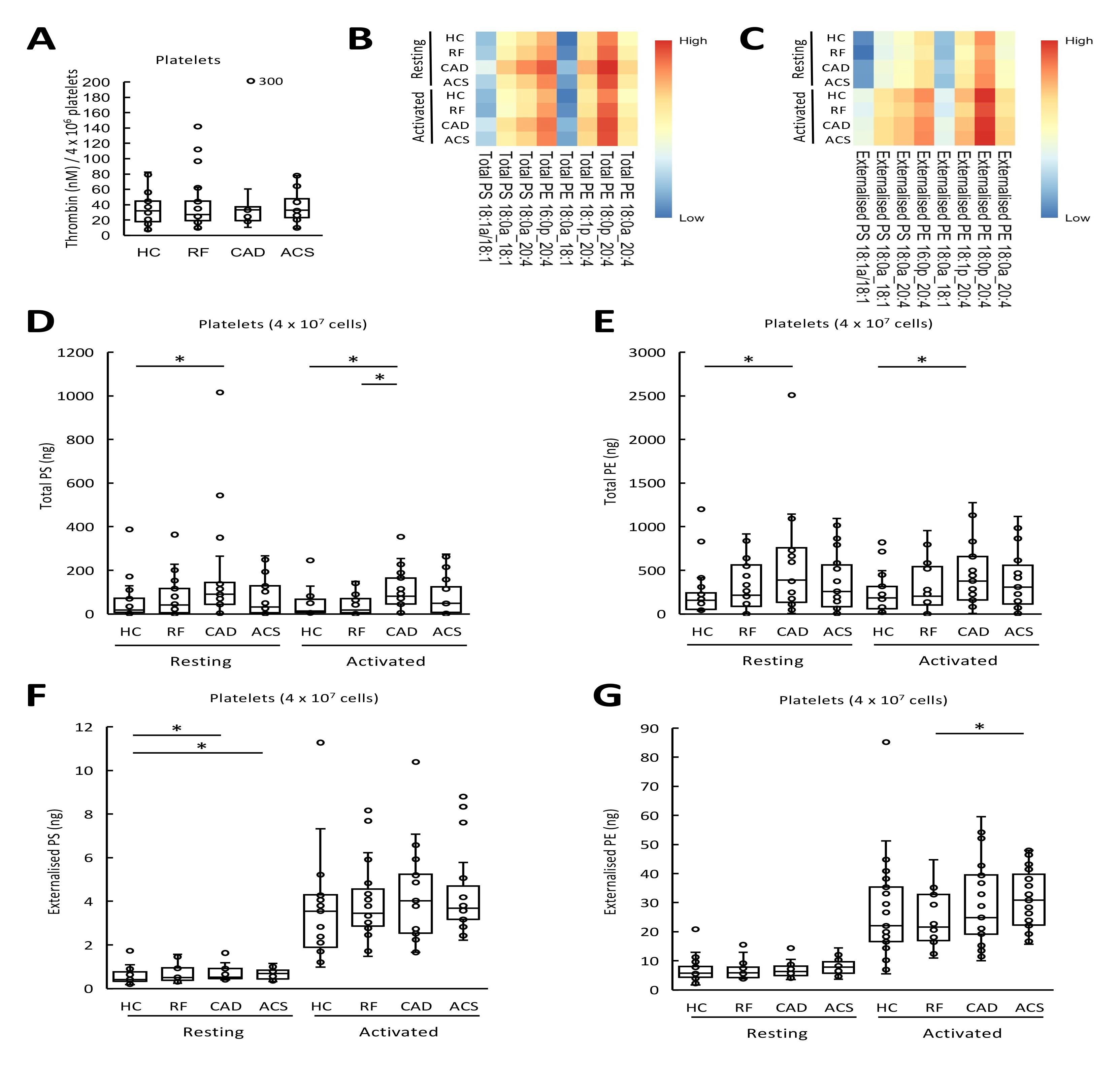
Thrombin generation on the surface of platelets in patients with arterial thrombosis was unchanged from HC, and there were minimal differences in aPL externalization between groups. *Panel A. Thrombin generation on the platelet surface was similar between clinical groups.* The ability of platelet membranes to support thrombin generation was assessed using the prothrombinase assay as in Methods and is displayed on a box plot. *Panels B,C. Heatmaps showing aPL molecular species in platelets.* Heatmaps were drawn using the pheatmap R package as described in Methods to visualize total (Panel B) and externalized (Panel C) aPL amounts between groups for all species measured using LC-MS/MS in resting or thrombin-activated platelets. *Panels D-G. Grouping by headgroup, the amounts of total PS, total PE, externalized PS and PE for each of the clinical groups demonstrates minimal differences between clinical groups.* Lipids were extracted from resting or thrombin-activated platelets as in Methods. Lipids amounts (ng) were determined using LC-MS/MS. Statistical significance was tested with Mann-Whitney-Wilcoxon test (*: p <0.05, **: p <0.01, ***: p <0.001). ACS: acute coronary syndrome (n=24), CAD: coronary artery disease but no ACS (n=19), RF: Risk factors with no significant coronary artery disease (n=23), HC: Healthy control (n=24).

### Platelets externalize aPL on activation, with an unchanged molecular composition between disease groups and healthy controls

Next, the pro-coagulant PL composition of platelets was determined using LC/MS/MS. The aPL species detected were as previously found in healthy control platelets, namely: PE 16:0p_20:4, PE 18:1p_20:4, PE 18:0a_20:4, PE 18:0p_20:4, PE 18:0a_18:1, PS 18:1a_18:1, PS 18:0a_18:1 and PS 18:0a_20:4, as previously shown^34^ (**Figure 3 B, Supplementary Figure 9**). Following thrombin activation, platelets from all groups externalized aPL with the most abundant isomers detected in higher amounts on the outside (p<0.001, **Figure 3 C**).

Total PS and PE levels were higher in platelets from the CAD group compared to HC both in resting and activated conditions (**Figure 3 D-E**). Following thrombin activation, externalized PS and PE rose compared to resting platelets but there was no significant differences between clinical groups and HC (**Figure 3 F-G**). Thus, disease did not result in major changes in either the amounts or molecular species of aPL externalized by platelets. Furthermore, there was no correlation between the amount of thrombin generated and externalized PS and PE amounts on the platelet membrane surface (**Supplementary Figure 8**).

In summary, platelets from patients with disease did not have higher PE/PS externalization or elevated thrombin generation capacity suggesting that changes in platelet aPL do not significantly contribute to the higher thrombotic risk seen in cardiovascular disease.

## Discussion

Thrombin generation on the plasma membrane surface relies on the presence of procoagulant lipids, primarily native PS and PE, but up to now it was not known whether thrombin generation or native aPL are altered in arterial thrombosis. This is important as it may influence the procoagulant status of patients with this condition and lead to further thrombotic complications^17, 18^. Previous studies have shown higher levels of plasma thrombin-anti thrombin complexes (TATs) in patients with a recent history of ACS, indicating activation of coagulation, however the mechanisms involved are unknown, in particular relating to the contribution of the pro-coagulant membrane ^19, 20^. Here, thrombin generation and the aPL composition of circulating blood cells and EV from patients with arterial vascular disease was characterized. Our data suggest that that EV and leucocyte membranes may contribute to increased thrombotic tendency of patients with cardiovascular disease. In contrast, a role for platelet aPL in driving membrane-dependent coagulation was not revealed. This has implications for the clinical management of ACS which currently does not target leukocyte or EV membrane compartments therapeutically for reducing thrombotic risk.

The assay employed in this study to assess thrombin generation is TF independent and no external PL was added. The means that the assay evaluates the role that the procoagulant membrane PL surface has on prothrombinase activity and is independent of changes in the levels of the patient’s coagulation factors. Whilst there were minimal differences in aPL amounts on the surface of EV and leukocytes between disease groups and HC as characterized by LC-MS/MS, the higher count of EV in plasma provided more surface area for the prothrombinase reaction to take place which in turn led to more thrombin generation (**Figure 1 A**). The same is likely for leukocytes which were found to be higher in patients with ACS compared with RF (**Supplementary Table 2**). Together, these results imply that a simple abundance of circulating procoagulant surfaces is sufficient to alter the amount of thrombin generation in resting states. Targeting EV clearance or shielding the circulation from the aPL membrane may therefore present a novel angle to reduce coagulation reactions in this thrombotic condition.

The use of LC-MS/MS to characterize aPL in platelets, leukocytes and EV has not been employed in arterial thrombosis patients prior to this study. Additionally, the external aPL lipidome in leukocytes and EV has not been previously characterized with LC-MS/MS. The majority of the literature on aPL trafficking and detection utilizes a flow cytometry-based method which relies on aPL-binding fluorescent probes^42–44^. The commonest of these is annexin V-FITC, which can bind to either PS or PE in the presence of calcium ^43, 44^. There are a number of limitations to this method, the main one being its non-quantitative nature. The binding of annexin V probes to cells and EV during flow cytometry exhibits rapidly saturated kinetics, likely as a result of steric hindrance preventing additional aPL from binding to this large protein^34^. Consequently, whilst it is feasible to count annexin V^+ve^ cells and EV using this method, it is not possible to quantify how much aPL is on the surface, distinguish between PS and PE or know what molecular species are exposed. This is relevant since recent studies have shown that the procoagulant activity of aPL is influenced by fatty acyl composition, with an impaired ability of PE comprising shorter FA chains to support coagulation ^34^. Mass spectrometry with biotin derivatization allows the quantitative analysis of aPL molecular species, distinguishing between external and total aPL amounts^41^. Our study provides a first look at the PS and PE species present on circulating membrane surfaces using an assay that distinguishes between total and externalized aPL^41^. The findings build on the current understanding of aPL externalization which had previously been reported in ACS patients using non-quantitative techniques with annexin V/lactadherin binding^45^.

Several studies have described higher numbers of EV in patients suffering from arterial thrombosis and its risk factors^46–52^. Hypertensive patients have more circulating EV which correlate with their blood pressure readings and are thought to be generated as a consequence of higher shear^46^. Similarly, diabetes mellitus is associated with higher levels of EV released as a consequence of stimuli such as advanced glycation products and oxidative stress, with a higher procoagulant phenotype in patients with poor glycemic control^47, 48^. Patients with dyslipidemias have upregulated EV levels due to LDL-induced membrane blebbing^49^. Indeed, the development of atherosclerosis may be influenced by EV, generated by mechanisms described above, which alter the profile of adhesion molecules on endothelial cells and promote monocyte transmigration and vascular inflammation^53, 54^. In addition, the presence of high amounts of EV within the atherosclerotic plaque may contribute to the thrombotic process that follows plaque rupture^55^. This may implicate EV in ACS where elevated numbers were demonstrated in comparison to CAD patients^50–52^, and shown to positively correlate with high-risk coronary angiographic features^56^. Nevertheless, beyond their proposed use as biomarkers and their elevated amounts, the exact role and regulation of EV in human arterial thrombosis remains largely unstudied.

Our study adds significantly to knowledge of EV biology in vascular disease, providing the first characterization of their aPL composition and how this contributes directly to thrombotic tendency. It is generally accepted that annexin V^+ve^ EV (containing externalized aPL) are procoagulant. Many studies have concluded that a higher number of EV implies more PS exposure in the circulation which in turn could lead to a more procoagulant phenotype^51, 57, 58^. This was observed in a study by Liu et al, where ACS patients were noted to have more annexin V^+ve^ EV compared with healthy controls^57^. In the same study, the authors demonstrated that *in-vitro* tenase and prothrombinase activity on EV-rich plasma fractions correlated positively with EV counts and was significantly reduced on blocking PS- binding sites with lactadherin^57^. However, methods used in the literature to study PS^+ve^ EV are limited to flow cytometry-based assays with annexin V. This marker is used to label EV with externalized aPL, but does not allow accurate quantification of external aPL, individual molecular species, nor does it determine the total aPL content within membranes. LC-MS/MS and biotin derivatization used in this paper overcomes these limitations, by mapping specific molecular species and their amounts, whilst also distinguishing between external and total membrane aPL ^41^.

The timing of any intervention targeting leukocytes or EV in ACS requires careful consideration. In this study, all ACS samples were taken within 48 h of the onset of the event, and therefore the findings reflect the acute phase. This immediate stage is associated with higher thrombotic risk^59^. As such, it is managed with more intensive anti-thrombotic treatment in the first few days after ACS such as the addition of a low molecular-weight heparin to dual anti-platelet therapy^59^. It is unclear however whether the higher procoagulant potential of leukocyte/EV membranes continues in the weeks or months after the acute event where persistent activation of the coagulation system (measured by higher plasma TATs) have been described in ACS^60^. To test this, longitudinal studies of patients with ACS in the acute (48 hours), intermediate (6 weeks) and distant (6 months) phase would provide more information on the variation of procoagulant membrane properties over time.

### Study Limitations

Whilst cells from patients with ACS may demonstrate lipidomic and biological differences, it is not possible to determine causality since it is not possible to predict when an ACS event will occur^61^. Furthermore, the EV cell of origin and TF surface content is unknown. Finally, the cross-sectional design of the study on the clinical cohort may be confounded by interindividual variations which may reduce statistical power. This, alongside the relatively small sample size, makes further validation and replication in a larger cohort advantageous.

## Conclusion

In this study, the procoagulant membrane in circulating blood cells and EV was determined in patients with arterial vascular disease. Our findings provide the first characterization of aPL in ACS circulating membranes and propose that the higher membrane procoagulant activity in arterial thrombosis is driven primarily by higher EV and leukocyte counts. Expanding on these findings, and testing whether interference with the procoagulant lipidome alters the observed incidence of arterial thrombosis would move the field closer towards targeting PL as therapeutic targets for the prevention of thrombosis in high risk groups.

## Supporting information

Data used for plotting figures

## Data Availability

All data produced in the present study are available upon reasonable request to the authors

---

**Supplementary Figure 1.**
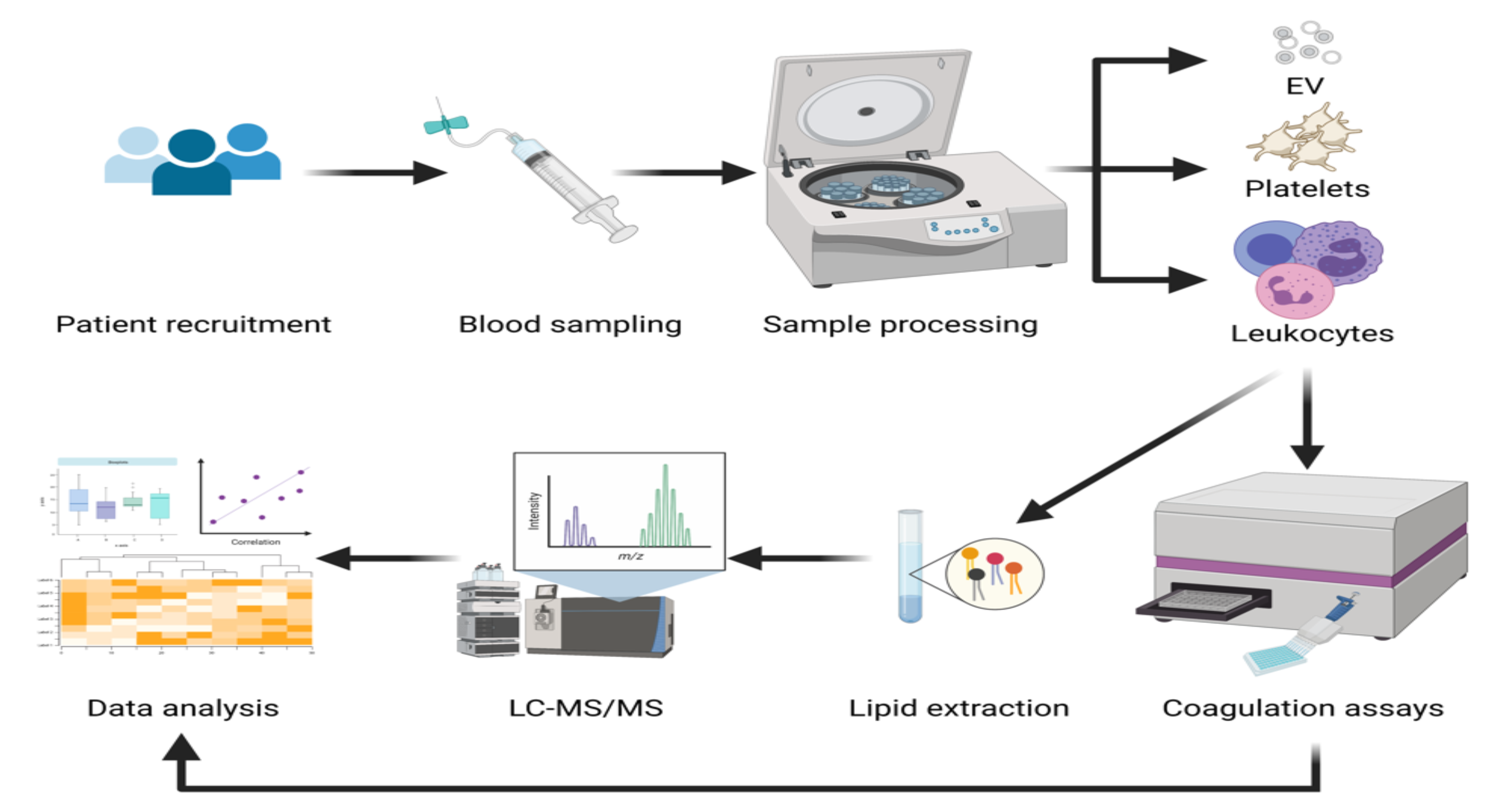
The experimental design of studies of arterial thrombosis in the clinical cohort. Patients were recruited as described in Methods. Blood was taken in a clinical area, and was transferred immediately to the laboratory. Platelets, leukocytes and extracellular vesicles (EV) were separated as described in Methods. The cells were divided into fractions to undergo functional testing with prothrombinase assay and lipid extraction and processing with LC-MS/MS, as outlined in Methods.

**Supplementary Figure 2.**
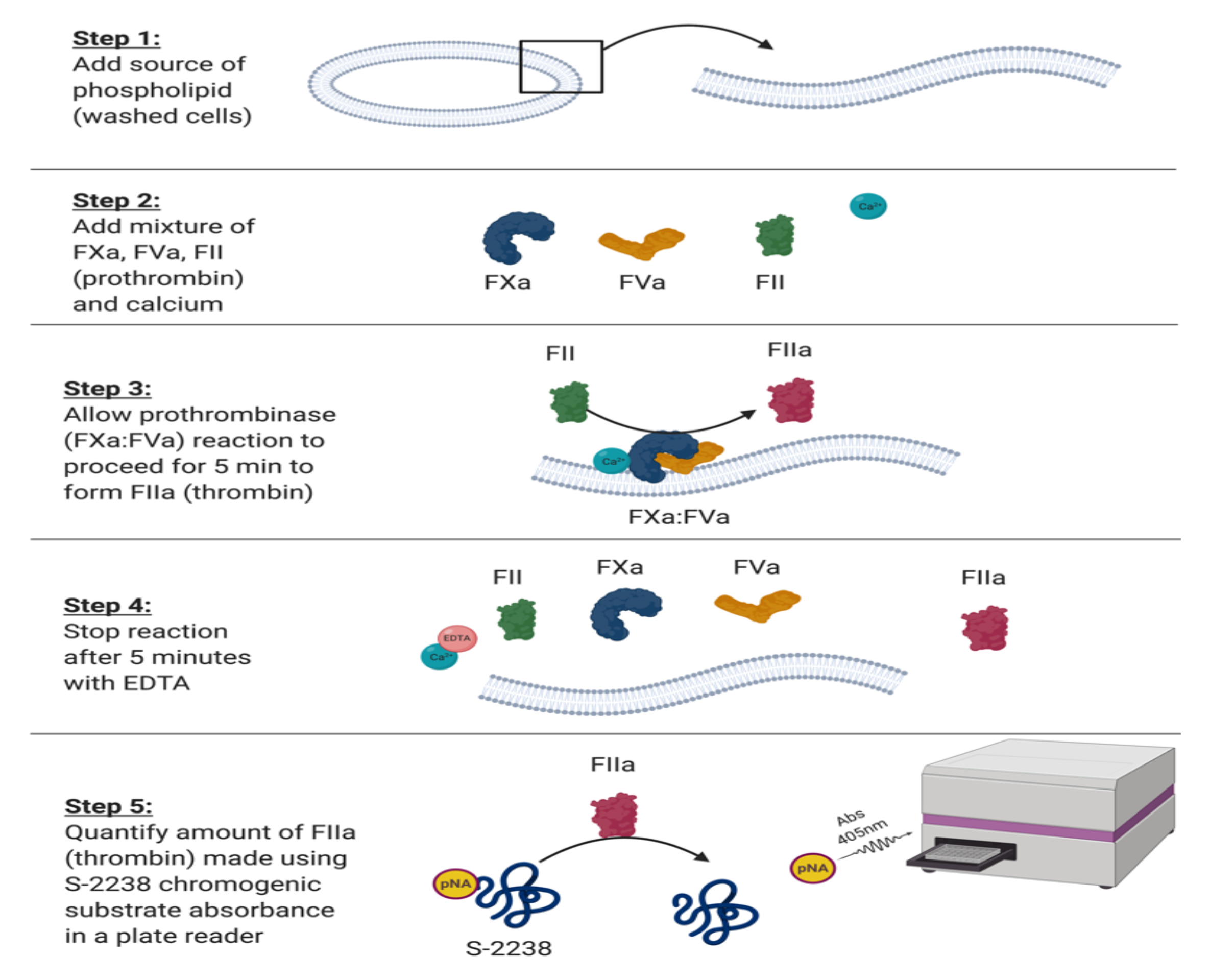
The workflow of the prothrombinase assay used to determine the procoagulant potential of washed cells and extra-cellular vesicles. In step 1, a known number of cells (platelets, leukocytes or EV) was aliquoted into a 96-well half-area plate. In step 2, a coagulation factor mixture containing calcium, FII, FXa, FVa was added to the plate to start the prothrombinase reaction on the surface of cells. The reaction was allowed to proceed for 5 min (step 3), before being quenched with EDTA (step 4). The amount of thrombin (FIIa) made was quantified using a p-nitroaniline (pNA) containing chromogenic substrate S- 2238 and absorbance read (405 nm) and compared to a standard curve of human thrombin (step 5).

**Supplementary Figure 3.**
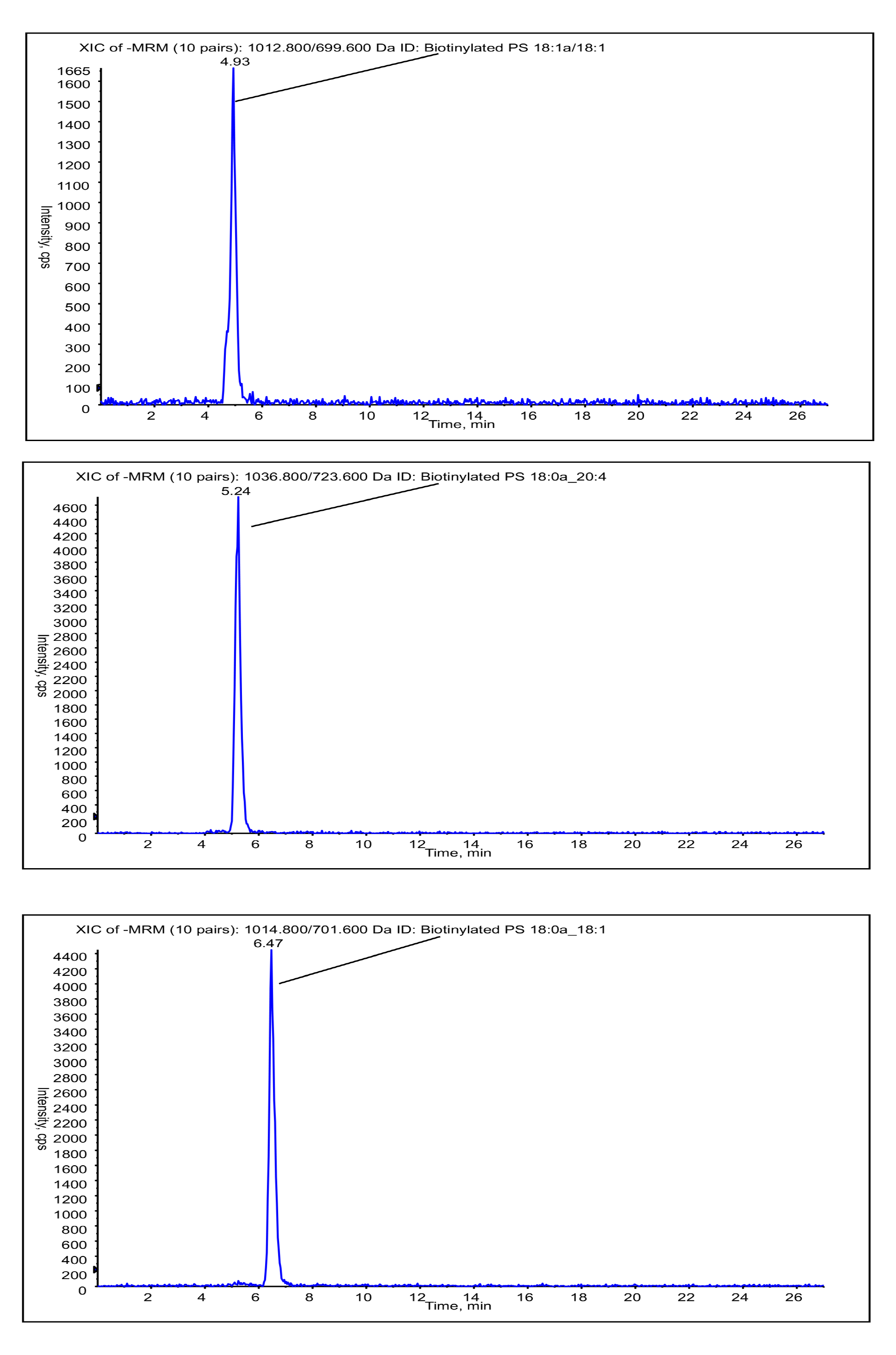
LC-MS/MS chromatograms of biotinylated-PS molecular species. Representative chromatograms from participant samples are shown for the PS molecular species quantified with the QTrap 4000 as in Methods. MRM transition and lipid name are displayed at the top of the chromatogram panels.

**Supplementary Figure 4.**
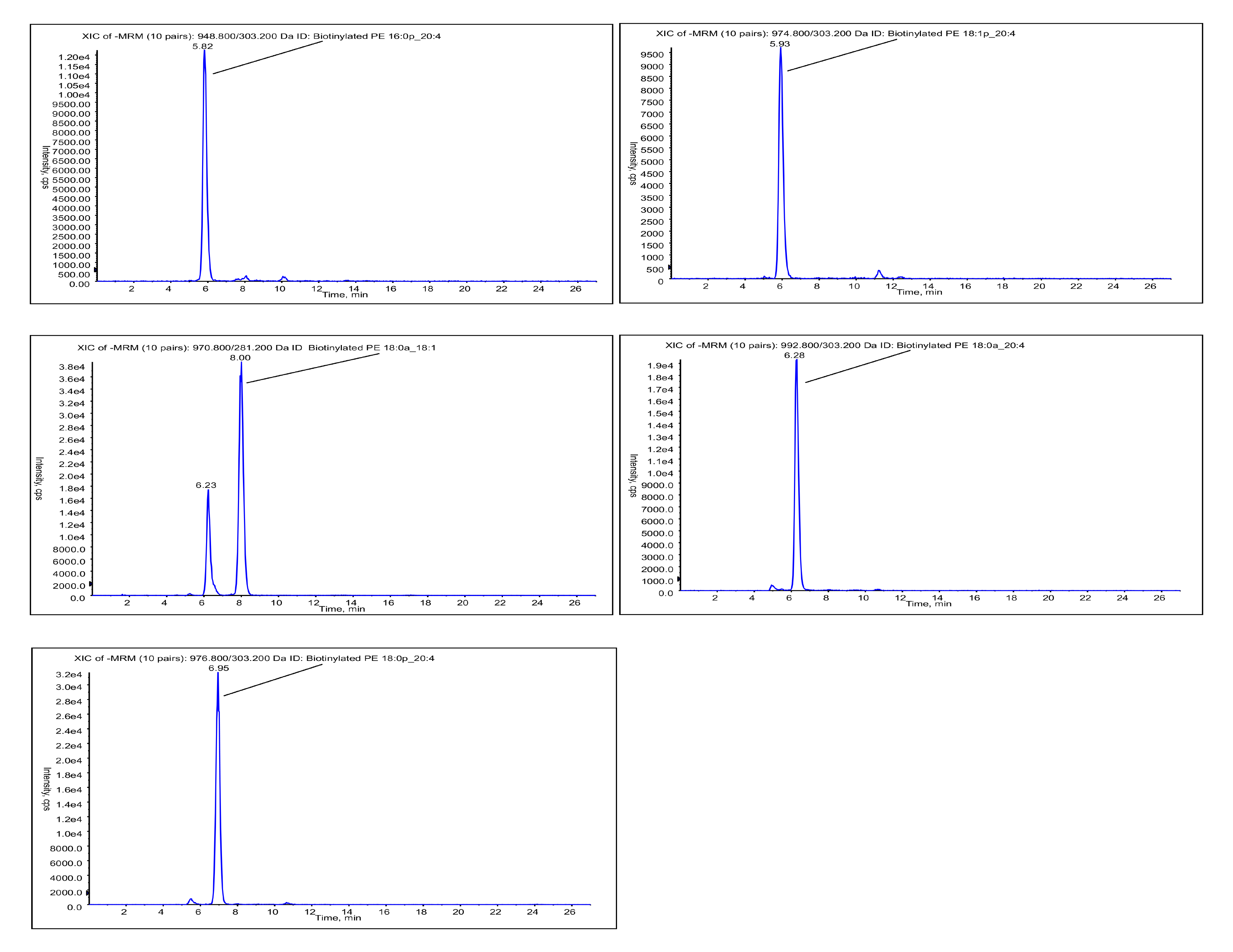
LC-MS/MS chromatograms of biotinylated-PE molecular species. Representative chromatograms from participant samples are shown for the PE molecular species quantified with the QTrap 4000 as in Methods. MRM transition and lipid name are displayed at the top of the chromatogram panels.

**Supplementary Figure 5.**
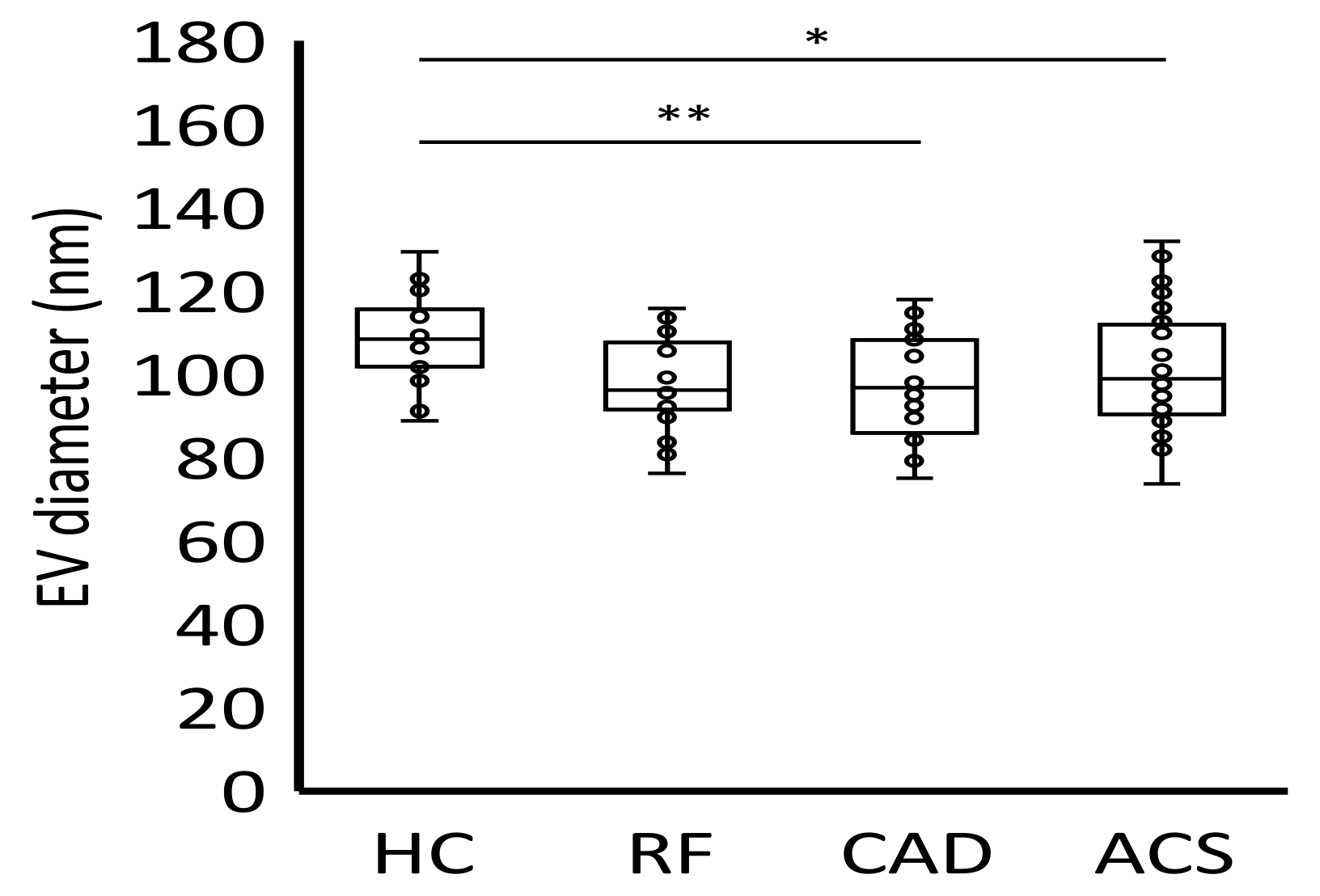
Significantly smaller EV particles (diameter) are detected in CAD patient plasma, and a downward trend in RF and ACS patients was seen, compared with healthy controls. Platelet-free plasma (0.5 ml) from each participant was processed through size exclusion chromatography (iZON qEV columns) and nanoparticle tracking analysis (Nanosight 300), as in Methods. Data for size was plotted as a box plot with the ggplot2 R package. Statistical significance was tested with Mann-Whitney-Wilcoxon test (*: p <0.05, **: p <0.01, ***: p <0.001). ACS: acute coronary syndrome (n=24), CAD: coronary artery disease but no ACS (n=19), RF: Risk factors with no significant coronary artery disease (n=23), HC: Healthy control (n=24).

**Supplementary Figure 6.**
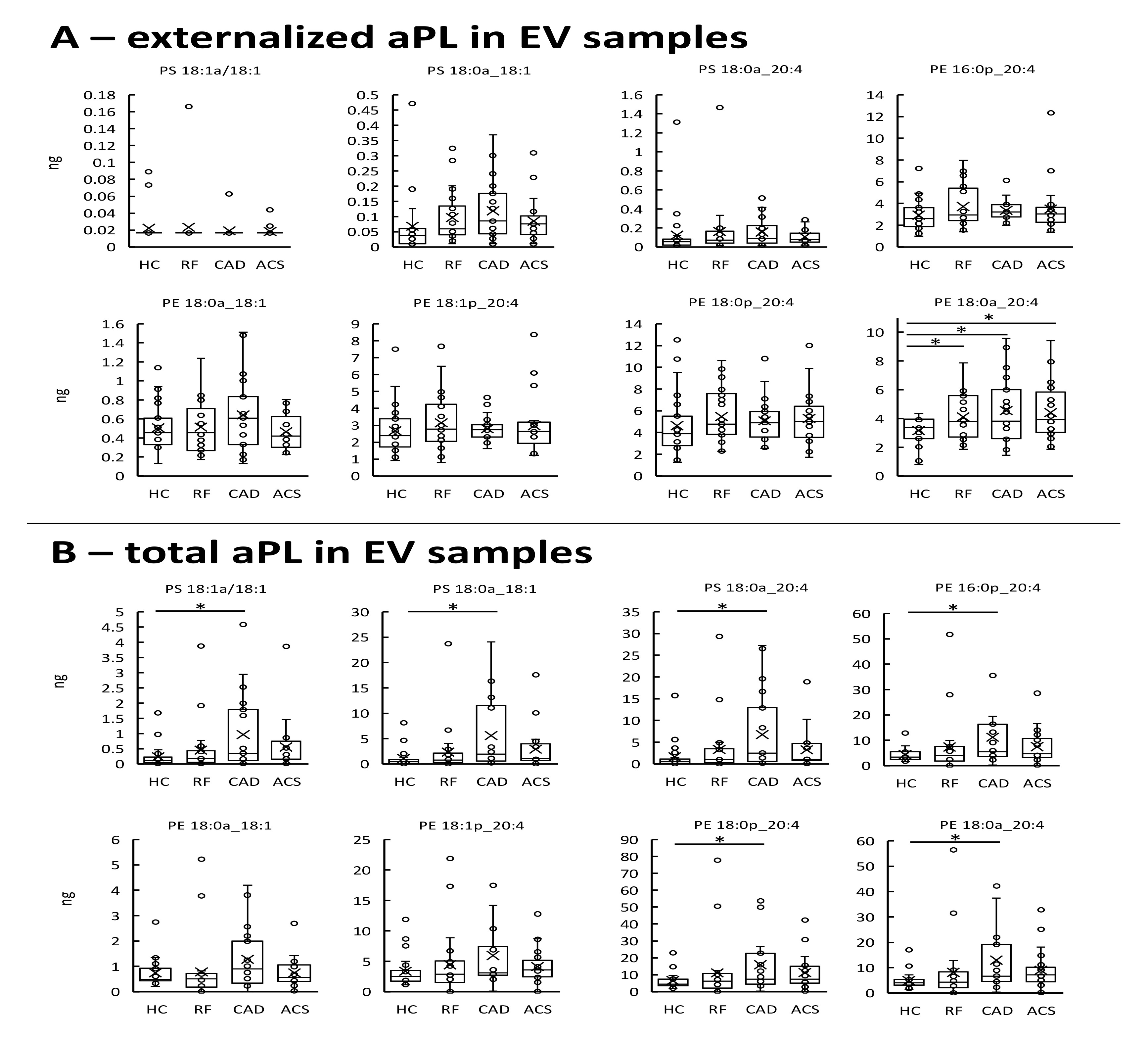
Characterization of aminophospholipid species in EV from a clinical cohort of patients with arterial thrombosis shows overall similar amounts of externalized aPL between clinical groups. *Panel A: The amounts of externalized aPL species in EV samples were largely similar between clinical groups.* Using EV samples isolated from plasma, the amounts of externalized PS and PE species for each of the clinical groups as quantified by LC-MS/MS were plotted to examine for differences between groups. *Panel B: The amounts of total aPL species in EV samples were elevated in the CAD group.* The amounts of total PS and PE species as quantified by LC-MS/MS for each of the clinical groups were plotted to examine for differences between groups. Lipids were extracted from EV-rich plasma fractions as in Methods. Lipid amounts (ng) were calculated by LC-MS/MS. Statistical significance was tested with heteroscedastic unpaired T-test (*: p <0.05, **: p <0.01, ***: p <0.001). ACS: acute coronary syndrome (n=24), CAD: coronary artery disease but no ACS (n=19), RF: Risk factors with no significant coronary artery disease (n=23), HC: Healthy control (n=24).

**Supplementary Figure 7.**
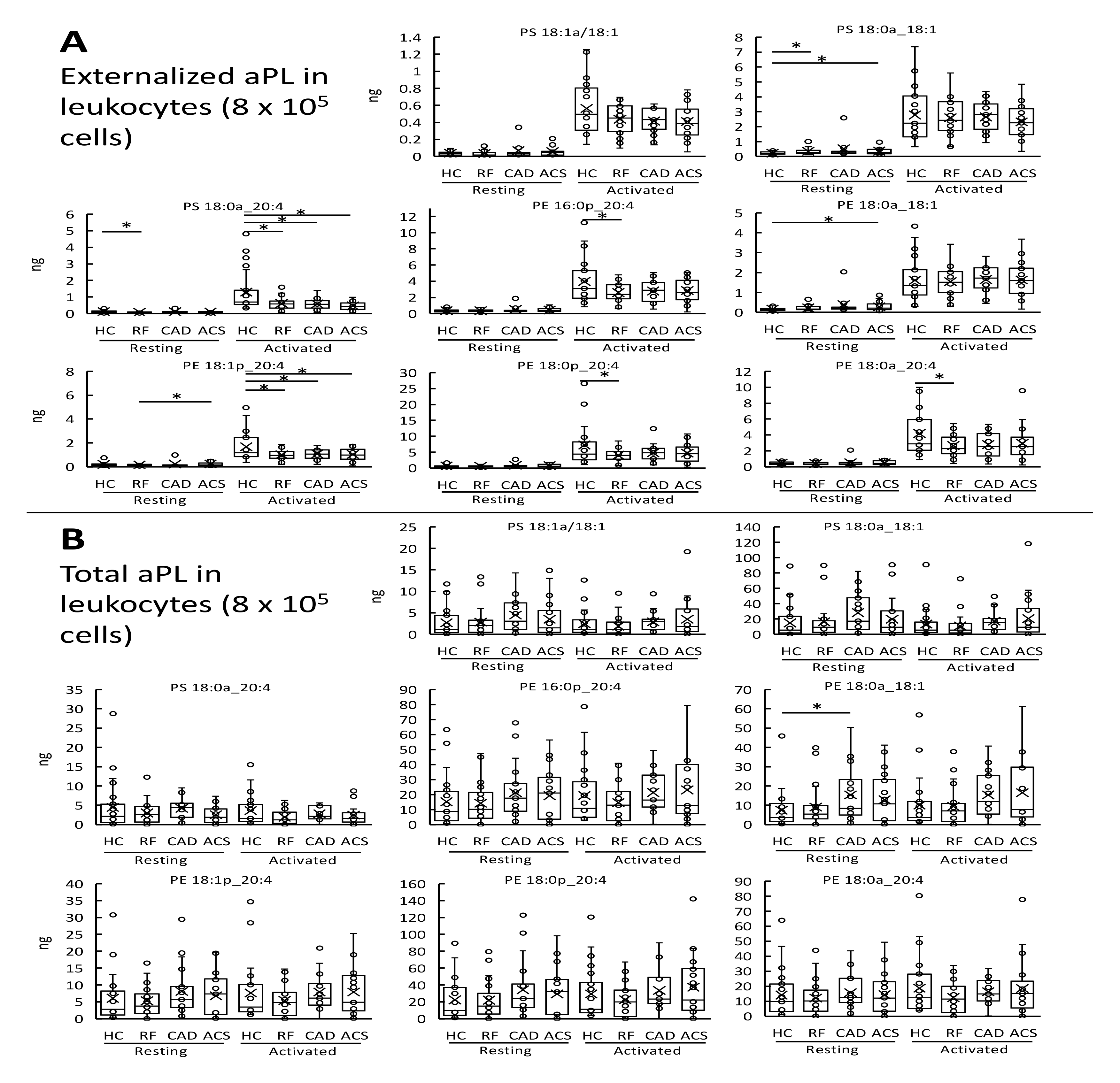
Characterization of aminophospholipid species in leukocytes from a clinical cohort of patients with arterial thrombosis shows largely similar profile of aPL lipids across clinical groups. *Panel A: Leukocytes externalize aPL molecular species upon ionophore-activation.* Using washed leukocytes (resting and ionophore activated), the amounts of externalized PS and PE species for each of the clinical groups as quantified by LC-MS/MS were plotted to examine for differences between groups. *Panel B: Minimal differences in the amounts of total aPL molecular species between clinical groups are seen.* The amounts of total PS and PE species for each of the clinical groups were quantified by LC-MS/MS to examine for differences between groups. Lipids were extracted from washed leukocytes as in Materials and Methods. Lipids amounts (ng) were calculated by LC-MS/MS. Statistical significance was tested with heteroscedastic unpaired T-test (*: p <0.05, **: p <0.01, ***: p <0.001). ACS: acute coronary syndrome (n=24), CAD: coronary artery disease but no ACS (n=19), RF: Risk factors with no significant coronary artery disease (n=23), HC: Healthy control (n=24).

**Supplementary Figure 8.**
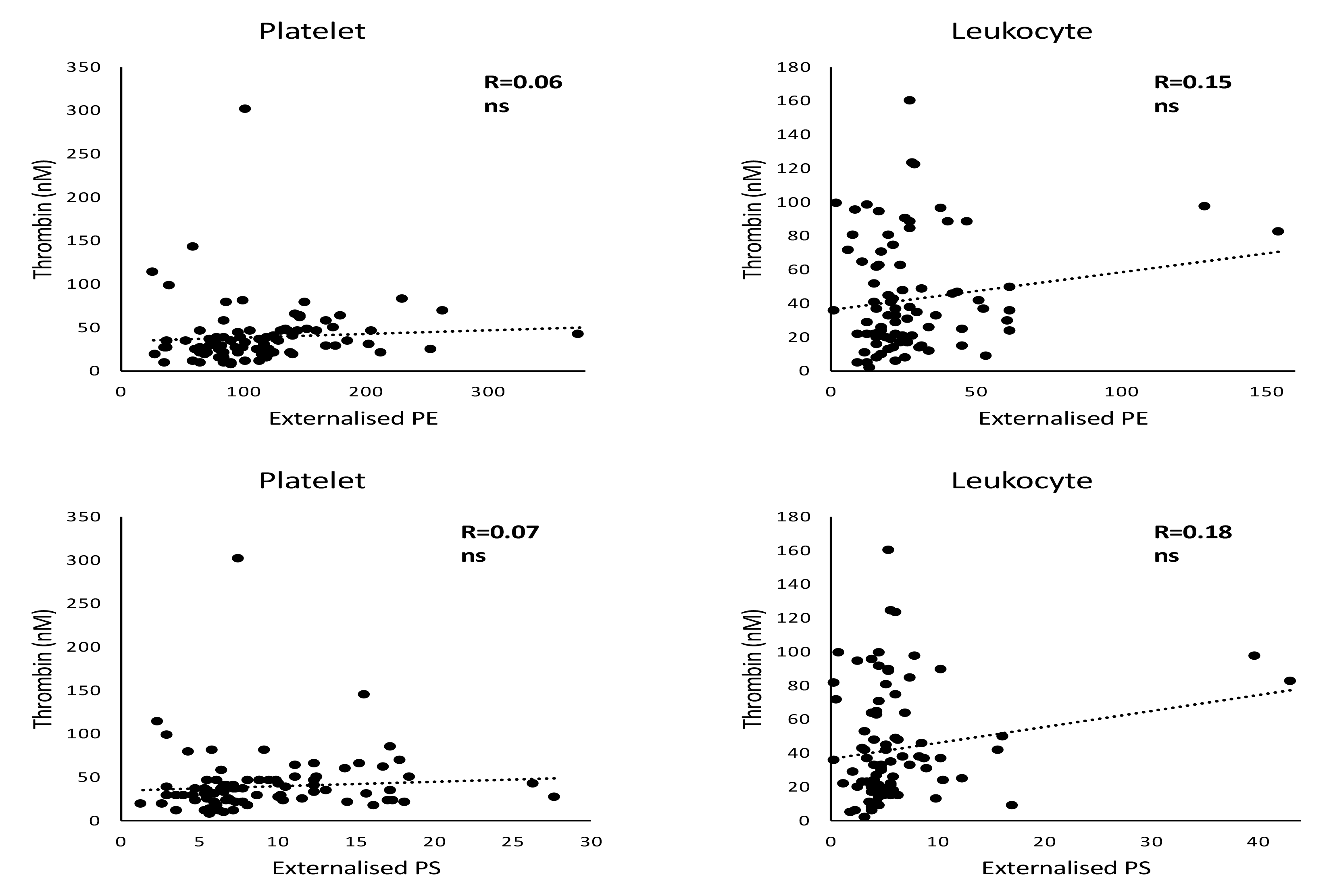
No correlation was seen between externalized aPL and thrombin generated on the surface of resting platelets and leukocytes. The ability of resting platelet and leukocyte membranes to support thrombin generation was assessed using the prothrombinase assay and correlated with externalized PS and PE amounts quantified by LC-MS/MS on using Pearson’s correlation. Lipids were extracted as in Materials and Methods. Lipids amounts (ng) were calculated by LC-MS/MS. Statistical significance was tested with Pearson’s correlation (n=90, the arterial thrombosis cohort described in Methods).

**Supplementary Figure 9:**
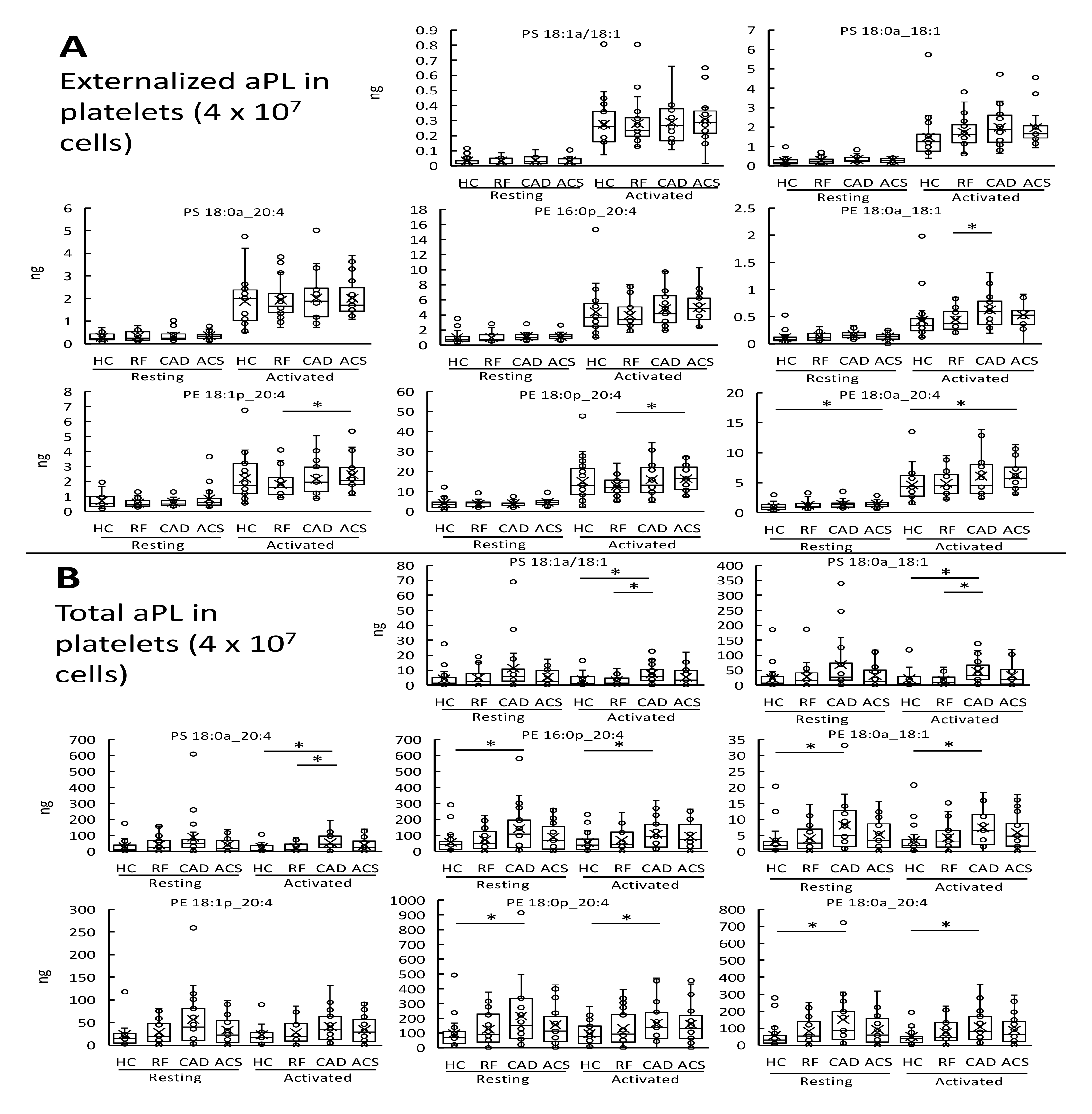
Characterization of aminophospholipid species in platelets from a clinical cohort of patients with arterial thrombosis shows a similar profile of aPL externalization between ACS and HC samples. *Panel A: Thrombin-activation increased aPL externalization on the surface of platelets with minimal differences between clinical groups.* Externalized PS and PE species were quantified in platelets (resting or thrombin-activated) by LC-MS/MS for each of the clinical groups, and plotted to examine for differences between groups. *Panel B: Quantification of Total aPL species detected in platelet membranes showed minimal differences between ACS and HC.* The amounts of total PS and PE for each clinical group was plotted to examine for differences between groups. Lipids were extracted from washed platelets as in Materials and Methods. Lipids amounts (ng) were calculated by LC-MS/MS. Statistical significance was tested with heteroscedastic unpaired T-test (*: p <0.05, **: p <0.01, ***: p <0.001). ACS: acute coronary syndrome (n=24), CAD: coronary artery disease but no ACS (n=19), RF: Risk factors with no significant coronary artery disease (n=23), HC: Healthy control (n=24).

**Supplementary Table 1.** Baseline clinical characteristics of patients recruited in the clinical cohort. (WCC: white cell count, RBC: red blood cell count, P2Y12 inhibitors: clopidogrel, prasugrel or ticagrelor, CKD: chronic kidney disease, SD: standard deviation, p-value tests: Fisher exact (categorical) or Kruskal-Wallis (continuous), p-value comparators: all clinical groups (age, gender) or all except HC for other variables)

| Variable | Healthy control<br>(HC)<br>(n=24) | Risk-factor matched<br>(RF)<br>(n=23) | Significant coronary artery<br>disease (CAD)<br>(n=19) | Acute coronary<br>syndrome (ACS)<br>(n=24) | p |
| --- | --- | --- | --- | --- | --- |
| Age, Mean ± SD | 64.92 ± 10.79 | 61.43 ± 8.16 | 64.53 ± 9.65 | 64.67 ± 9.88 | 0.473 |
| Male Gender (%) | 14 (58.3) | 12 (52.2) | 16 (84.2) | 17 (70.8) | 0.133 |
| Creatinine μmol/L, Mean ± SD | - | 77.57 ± 12.60 | 88.39 ± 31.95 | 84.79 ± 18.64 | 0.318 |
| Haemoglobin g/dL, Mean ± SD | - | 146.04 ± 13.03 | 138.24 ± 17.95 | 144.46 ± 18.88 | 0.380 |
| Platelets $\times 10^9$/L, Mean ± SD | - | 250.26 ± 44.01 | 261.94 ± 56.08 | 271.50 ± 82.72 | 0.869 |
| WCC $\times 10^9$/L, Mean ± SD | - | 6.96 ± 1.57 | 7.92 ± 1.92 | 9.28 ± 2.90 | 0.011 |
| Neutrophils | - | 4.33 ± 1.17 | 5.02 ± 1.32 | 6.51 ± 2.49 | 0.003 |
| Eosinophils | - | 0.19 ± 0.09 | 0.25 ± 0.12 | 0.11 ± 0.08 | <0.001 |
| Basophils | - | 0.02 ± 0.04 | 0.03 ± 0.05 | 0.00 ± 0.02 | 0.098 |
| Lymphocytes | - | 1.82 ± 0.68 | 1.89 ± 0.70 | 1.85 ± 0.71 | 0.818 |
| Monocytes | - | 0.57 ± 0.19 | 0.68 ± 0.21 | 0.73 ± 0.32 | 0.150 |
| RBC $\times 10^{12}$/L, Mean ± SD | - | 4.80 ± 0.42 | 4.14 ± 1.26 | 4.64 ± 0.54 | 0.090 |
| Aspirin use (%) | 0 (0) | 20 (87) | 14 (73.7) | 24 (100) | 0.017 |
| P2Y12 inhibitor use (%) | 0 (0) | 3 (13) | 6 (31.6) | 24 (100) | <0.001 |
| Anticoagulant use (%) | 0 (0) | 3 (13) | 0 (0) | 24 (100) | - |
| Statin use (%) | 0 (0) | 15 (65.2) | 15 (78.9) | 19 (79.2) | 0.532 |
| Hypertension (%) | 0 (0) | 13 (56.5) | 13 (68.4) | 11 (45.8) | 0.350 |
| Diabetes (%) | 0 (0) | 7 (30.4) | 3 (15.8) | 6 (25) | 0.554 |
| Smoker (%) | 0 (0) | 7 (30.4) | 9 (47.4) | 13 (54.2) | 0.244 |
| CKD (%) | 0 (0) | 0 (0) | 3 (15.8) | 3 (12.5) | - |

**Supplementary Table 2.**
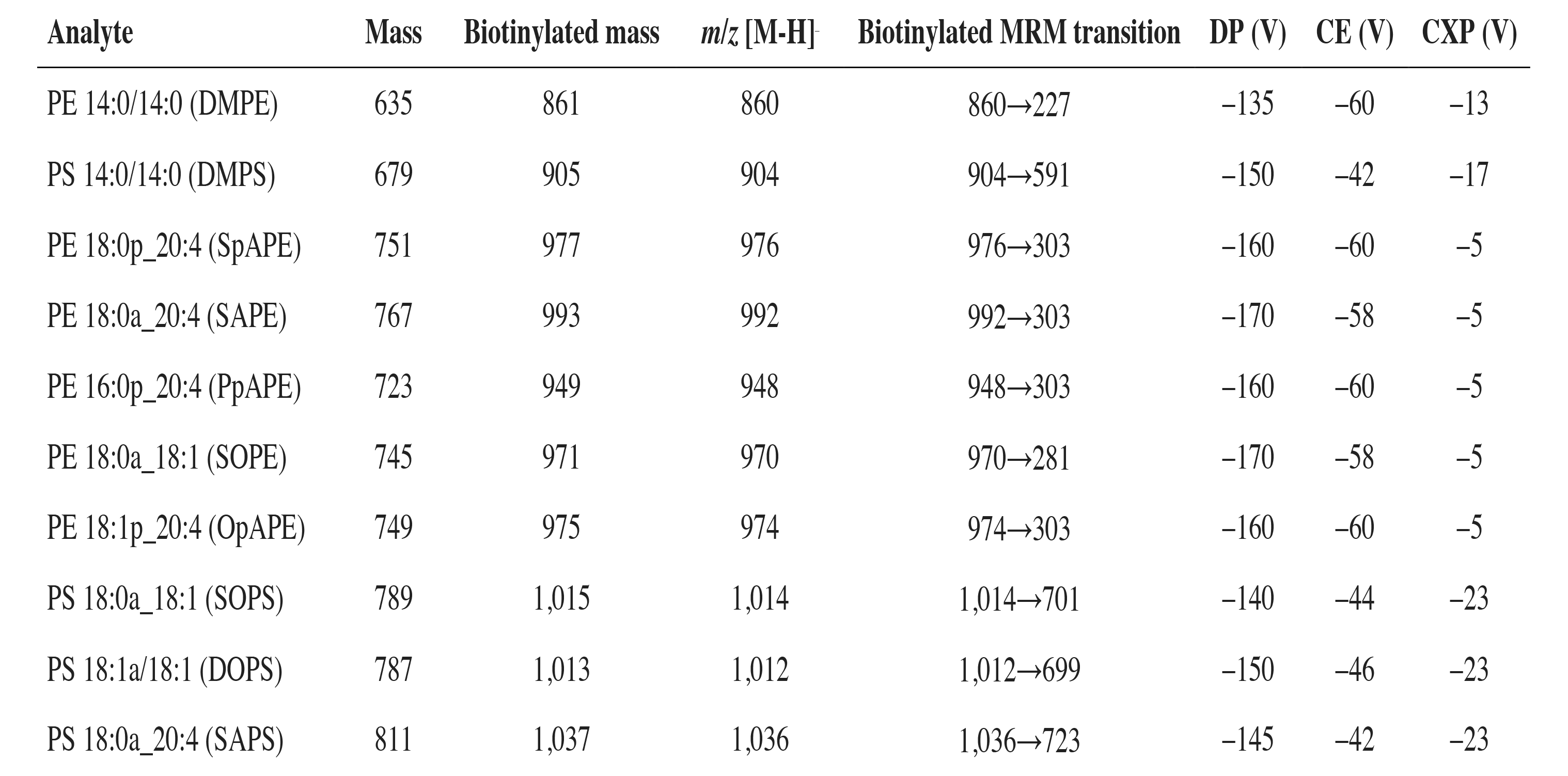
**Multiple reaction monitoring (MRM) transitions for the aPL analyzed in negative ion mode as part of targeted lipidomic analysis of cells isolated from patients with atherothrombosis.**

| Analyte | Mass | Biotinylated mass | *m/z* [M-H]<sup>-</sup> | Biotinylated MRM transition | DP (V) | CE (V) | CXP (V) |
| --- | --- | --- | --- | --- | --- | --- | --- |
| PE 14:0/14:0 (DMPE) | 635 | 861 | 860 | 860→227 | -135 | -60 | -13 |
| PS 14:0/14:0 (DMPS) | 679 | 905 | 904 | 904→591 | -150 | -42 | -17 |
| PE 18:0p_20:4 (SpAPE) | 751 | 977 | 976 | 976→303 | -160 | -60 | -5 |
| PE 18:0a_20:4 (SAPE) | 767 | 993 | 992 | 992→303 | -170 | -58 | -5 |
| PE 16:0p_20:4 (PpAPE) | 723 | 949 | 948 | 948→303 | -160 | -60 | -5 |
| PE 18:0a_18:1 (SOPE) | 745 | 971 | 970 | 970→281 | -170 | -58 | -5 |
| PE 18:1p_20:4 (OpAPE) | 749 | 975 | 974 | 974→303 | -160 | -60 | -5 |
| PS 18:0a_18:1 (SOPS) | 789 | 1,015 | 1,014 | 1,014→701 | -140 | -44 | -23 |
| PS 18:1a/18:1 (DOPS) | 787 | 1,013 | 1,012 | 1,012→699 | -150 | -46 | -23 |
| PS 18:0a_20:4 (SAPS) | 811 | 1,037 | 1,036 | 1,036→723 | -145 | -42 | -23 |

